# Childhood Trauma and Trajectories of Depressive Symptoms Across Adolescence

**DOI:** 10.1101/19002519

**Authors:** Alex S. F. Kwong, Jennifer M. Maddalena, Jazz Croft, Jon Heron, George Leckie

## Abstract

**Background:** Growth curve modelling such as trajectory analysis is useful for examining the longitudinal nature of depressive symptoms, their antecedents and later consequences. However, issues in interpretation associated with this methodology could hinder the translation from results to policy changes and interventions. The aim of this article is to provide a “model interpretation framework” for highlighting growth curve results in a more interpretable manner. Here we demonstrate the association between childhood trauma and trajectories of depressive symptoms. Childhood trauma has been shown to a be strong predictor for later depression, but less is known how childhood trauma has an effect throughout adolescence and young adulthood. Identifying when childhood trauma (and its severity) is likely to have its greatest impact on depression is important for determining the timing of interventions for depression.

**Methods:** We used data on over 6,500 individuals from the Avon Longitudinal Study of Parents and Children (ALSPAC) to estimate trajectories of depressive symptoms between the ages of 11 and 24. Depressive symptoms were measured using the short mood and feelings questionnaire (SMFQ) across 9 occasions. Childhood trauma was assessed between the ages of 5 and 10 years old, and we estimated population averaged multilevel growth curves of depressive symptoms for exposure to trauma (yes vs no) and then in a separate model, the number of trauma types reported such as inter-personal violence or neglect (coded as 0, 1, 2, 3+). We then calculated what the depressive symptoms scores would be ages 12, 14, 16, 18, 20, 22, 24, between these varying trajectories.

**Results:** Reported exposure to childhood trauma was associated with less favourable trajectories of depressive symptoms across adolescence, mainly characterised by exposed individuals having worse depressive symptoms at age 16. There was an exposure-response relationship between the number of childhood traumas and trajectories of depressive symptoms.

Individuals exposed to 3 or more types of trauma had substantially steeper and less favourable trajectories of depressive symptoms: becoming worse at a more rapid rate until the age of 18. By age 18, individuals that reported the greatest exposure to trauma (3+ types of trauma) had 14% more depressive symptoms compared to non-exposed participants.

**Limitations:** This study was subject to attrition, particularly towards the latter ages of the SMFQ.

**Conclusion:** Childhood trauma is strongly associated with less favourable trajectories of depressive symptoms across adolescence. Individuals exposed to multiple types of inter-personal violence or neglect are at the greatest risk of worsening depressive symptoms throughout adolescence and young adulthood. Individuals exposed to traumatic experiences in childhood should be identified as at high risk of depression and other adverse outcomes as early trauma may disrupt social development and have lasting consequences on mental health outcomes.

The model interpretation framework presented here may be more interpretable for researchers, clinicians and policy makers as it allows comparisons of depression across multiple stages of development to highlight when the effects of depression are greatest.

## 1. Introduction

Growth curve modelling is useful for examining the longitudinal nature of depression across development (Musliner, Munk-Olsen, Eaton, & Zandi, 2016; Sutin et al., 2013). Growth curve models allow researchers to estimate trajectories that describe patterns of change in depressive mood over time. These are potentially more useful than analysis of data from any single occasion, as it is possible to estimate the longitudinal nature of depression across periods of development. Furthermore, it is possible to examine the antecedents of varying trajectories of depressive symptoms (Ferro, Gorter, & Boyle, 2015a; Ge, Natsuaki, & Conger, 2006; Mazza, Fleming, Abbott, Haggerty, & Catalano, 2010), their later consequences (Whalen et al., 2016; Yaroslavsky, Pettit, Lewinsohn, Seeley, & Roberts, 2013), their relationship with other co-occurring traits (Marmorstein, 2009; Mumford, Liu, Hair, & Yu, 2013; Wiesner & Kim, 2006) and the identification of critical points that could be translated into preventions and interventions (Kwong et al., 2019). Consequently, growth curve or trajectory analysis is not only a useful tool for research, but also potentially for clinical inference and identifying at risk groups for treatment (Schubert, Clark, Van, Collinson, & Baune, 2017).

Methodology exploring trajectories of depressive symptoms has been increasingly used in recent years (Musliner et al., 2016; Schubert et al., 2017; Shore, Toumbourou, Lewis, & Kremer, 2018). However, much of this research has yet to impact on policy and clinical inference, which could be a result of difficulties in translating findings to inference. Estimating trajectories of depressive symptoms can be done in several ways: one approach is to plot the mean scores by age and depressive symptoms and compare differences at each age (i.e., with a t-test/ANOVA). However, one problem with this approach is that there may be different samples at the various ages, and this can lead to biased estimates with missing data. A more popular approach is to stratify population trajectories into multiple sub-group trajectories (e.g., latent classes growth analysis or growth mixture models, see (Muthen & Muthen, 2000)). An alternative approach is to estimate population-averaged trajectories, with individuals being allowed to vary around this population average (e.g., multilevel growth-curves and structural equation models, see (Hedeker & Gibbons, 2006)). These latter approaches quantify the degree of change, making them useful methods for estimating how depression changes over a certain period of time, and what might predict this change. These two approaches are somewhat similar but can produce different results and interpretations, thus harmonising results from both analyses is challenging and could impact on how clinicians and policy makers interpret and implement these findings.

One issue with the longitudinal analyses of depressive mood is that depression fluctuates and is characterised by periods of rapid growth and varying ages of onset (Ferro, Gorter, & Boyle, 2015b; Kwong et al., 2019), especially during heightened risk periods such as adolescence (Thapar, Collinshaw, Pine, & Thapar, 2012). In many studies, modelling trajectories of depressive symptoms and the subsequent interpretation is complex. For example, changes in depression are rarely linear. Studies may typically use higher order polynomials to model non-linear trajectories (e.g., quadratic and cubic functions of age), which can result in complicated model estimates (positive quadratic terms but negative cubic terms etc). This can be difficult for researchers and clinicians to interpret and inform policy and practise. Therefore, it is important to find alternative ways to highlight results from trajectory methods. An example of this has been demonstrated in longitudinal assessments of autistics traits (Mandy, Pellicano, St Pourcain, Skuse, & Heron, 2018). Here, Mandy and colleagues estimated differences in the predicted autistic traits scores between male and female trajectories at several ages.

This approach could be beneficial when applied to the longitudinal study of depressive symptoms. It is often important to not only highlight the course of depression over time, but also when differences may be occurring and whether two or more trajectories (i.e., exposed to trauma vs not exposed to trauma) differ at a certain age or stage of development. The results in many studies highlighting differences between trajectories are not straight forward, as the presented estimates are typically the estimated coefficients of polynomials and the coefficient of each term in the polynomial is hard to interpret in isolation. More clarity in this area could assist clinicians and policy makers to understand the implications of results from analyses of trajectories of health outcomes. Therefore, the first aim of this study is to highlight how the results from these analyses can be made more interpretable to researchers, clinicians and policy makers by presenting an alternative model interpretation framework to highlight results that show the depressive symptoms scores of different trajectories at different ages.

One of the most useful applications for growth curve modelling is to examine how early antecedents or risk factors may predict the trajectory of depressive symptoms over time. Here, we use the example of exposure to childhood trauma to examine its association with depressive symptoms across adolescence and young adulthood. Childhood traumas is a key predictor for adolescent and young adulthood depression (Copeland et al., 2018; Lewis et al., 2019; Sara & Lappin, 2017), yet its effect on depression throughout the course of adolescence and young adulthood is still not clear.

Latent class analysis has found that exposure to abuse before the age of 18 years old is associated with less favourable trajectories of depressive symptoms (Olino, Klein, Lewinsohn, Rohde, & Seeley, 2010). Here, Olino and colleagues found that individuals who scored higher on a scale of childhood adversity and abuse has increased odds of belonging to the ‘persistent’, ‘increasing’ and ‘initially high’ trajectories, but interestingly not the ‘later onset’ trajectory of depressive symptoms. Maltreatment in childhood has also been associated with steeper trajectories of depressive symptoms, and those who experienced sexual abuse younger were more likely to have trajectories that started off higher, but then reduced over time (Carlson & Oshri, 2018; Keane, Magee, & Kelly, 2018). These results suggest that the effects of childhood trauma may have more damaging effects during the formative years of adolescence, as opposed to an increased risk of depression at a later age. There is also evidence to suggest that more stressful life events in childhood are associated with higher trajectories of depressive symptoms in other longitudinal studies (Fernandez Castelao & Kroner-Herwig, 2013; Lee, Wickrama, Kwon, Lorenz, & Oshri, 2017; Stoolmiller, Kim, & Capaldi, 2005; Weeks et al., 2014). However, in many of these studies, depressive symptoms are not measured across the entire period of adolescence and therefore the true longitudinal nature of childhood trauma on trajectories of depressive symptoms is not known.

For population averaged trajectories, there is less evidence that negative life events in childhood may impact on trajectories of depressive symptoms. However, recent studies have found that adverse childhood experiences including abuse and neglect were associated with less favourable trajectories of depressive symptoms (Barboza & Dominguez, 2017) and that increased stressful life events were associated with a steeper slope for trajectories of depressive symptoms across adolescence (Adkins, Wang, Dupre, van den Oord, & Elder, 2009; Ge, Lorenz, Conger, Elder, & Simons, 1994).

Identifying when childhood trauma is likely to have its greatest impact on depression throughout adolescence and young adulthood is important for indicating when interventions and support may be most beneficial. Currently, there is a paucity of knowledge on when childhood trauma is likely to have the greatest impact on trajectories of depressive symptoms. There is also some evidence to suggest that negative life events such as trauma and stressful life events are not associated with worsening trajectories (Mazza et al., 2010; Sallinen, Rönkä, Kinnunen, & Kokko, 2016). However, we note that in one of these studies, negative life events were assessed by parental report (Mazza et al., 2010), which could lead to an underreporting of the exposure.

Research has also yet to focus on how the number of traumatic experiences may predict the longitudinal nature of depression. There is some evidence to suggest that the number of stressful life events may explain the extent to which the risk of depression is increased by exposure to trauma. More than two stressful life events early in childhood have been associated with higher trajectories of depressive symptoms (Weeks et al., 2014).

However, little is known about traumatic experiences in childhood and later trajectories of depression. Life-course models suggest that an accumulation of negative events may be more harmful than traumatic exposure on a single occasion (Dunn, Soare, et al., 2018). It is therefore important to examine if the number of childhood traumatic experiences might differentially predict varying trajectories of depressive symptoms across adolescence.

Thus, the second aim of the current study is to build upon previous research by examining the association between childhood trauma and trajectories of depressive symptoms across adolescence and young adulthood. We are also interested in extending previous research by investigating how this association might change depending on the number of childhood traumatic experiences.

## 2. Method

### 2.1. Study sample

We used data from the Avon Longitudinal Study of Parents and Children (ALSPAC), a birth cohort that recruited pregnant women residing in Avon, UK with expected dates of delivery 1^st^ April 1991 to 31^st^ December 1992 (Boyd et al., 2013; Fraser et al., 2013). The initial cohort consisted of 14,062 children and all participants provided written informed consent. Ethical approval was obtained from the ALSPAC Ethics and Law Committee and the Local Research Ethics Committees. The study website contains details of all the data that is available through a fully searchable data dictionary and variable search tool: http://www.bristol.ac.uk/alspac/researchers/our-data. Part of this data was collected using REDCap, please see the REDCap website for details (https://projectredcap.org/resources/citations/).

### 2.2. Measures

#### 2.2.1. Depressive symptoms

Self-reported depressive symptoms were measured on nine occasions between ages 10 and 24 using the short mood and feelings questionnaire (SMFQ) (Angold, Costello, Messer, & Pickles, 1995). The SMFQ is a 13-item questionnaire that measures the presence of depressive symptoms in the last two weeks and was administered via postal questionnaire or in clinics. Each item is scored 0,1 or 2, resulting in a summed score between 0-26. The SMFQ correlates strongly with clinical depression (Thapar & McGuffin, 1998; Turner, Joinson, Peters, Wiles, & Lewis, 2014) and has been used to explore trajectories of depressive symptoms in other studies (Kwong et al., 2019; Mahedy et al., 2017; Rice et al., 2018).

#### 2.2.2. Trauma

Two measures of trauma were used for this analysis derived from previous analysis (Croft et al., 2018; Houtepen, Heron, Suderman, Tilling, & Howe, 2018): (1) A binary measure of inter-personal violence and neglect during mid-childhood (age 5-10.9 years; [no=0; yes=1]) and (2) the number of trauma types reported during mid-childhood (0, 1, 2 or 3 or more [3+]). Briefly, these variables were derived from 121 self-report and parent report questions about traumatic experiences. The data used to create these variables were taken from multiple trauma questions collected from both parents and children regarding exposure to sexual abuse, physical abuse, emotional abuse, emotional neglect, domestic violence and bullying.

#### 2.2.3. Confounders

The following confounders were included based upon previous literature examining early social risk factors and trajectories of depressive symptoms (Kingsbury et al., 2016; Kwong et al., 2019; Mahedy et al., 2017): sex (coded as a dummy variable for being female [male = 0; female =1]), maternal postnatal depression (no vs yes), maternal socioeconomic status at birth (manual vs non-manual), maternal educational attainment at birth (A-level or higher vs O-level vs < O-level) and parity (1^st^ born vs 2^nd^ born vs 3^rd^ born or more).

### 2.3. Statistical methods

Trajectories of depressive symptoms were estimated using multilevel growth-curve modelling (Hedeker & Gibbons, 2006; Raudenbush & Bryk, 2002). Briefly, multilevel growth-curve models estimate population averaged trajectories, with individual level trajectories varying around these population averages (i.e., each person can have their own trajectory that deviates from the average for their population). Descriptive statistics and previous analysis on this data has shown that change in depressive symptoms over time is non-linear (Kwong et al., 2019). To model this non-linearity, a multilevel quartic age polynomial growth-curve model was chosen in line with previous research using higher-order age polynomials for examining trajectories of depressive symptoms (Ferro et al., 2015a; Kwong et al., 2019; Rawana & Morgan, 2014). Age was first grand-mean centred around 16.53 years (the mean age of all assessments) in order to improve interpretation (Ferro et al., 2015a; Kwong et al., 2019; Rawana & Morgan, 2014), since model intercept and intercept variance then correspond to the middle of adolescence. Model fit was assessed using likelihood-ratio tests and information criteria, consistent with other studies using multilevel growth-curve models (Singer & Willett, 2003).

We ran separate models to examine the association between each trauma variable and these trajectories of depressive symptoms. The first model examined population-averaged trajectories for the exposure to trauma (no vs yes) by interacting the fixed-effects quartic age polynomial terms with the “any trauma” dummy variable. This produced two trajectories of depressive symptoms corresponding to the sub-populations with and without trauma. The second model repeated this approach by entering dummy variables representing the number of traumatic experiences (1, 2 and 3+). In all the analyses, the quartic polynomial age terms varied randomly across individuals to capture each individual’s unique trajectory.

We then predicted depressive symptoms scores at each of the following ages: 12, 14, 16 18, 20, 22 and 24, separately for each population averaged trajectory. We used the delta method to compare these predicted scores across the different trajectories at each of these ages.

All analyses were conducted using Stata 15 (StataCorp, College Station, TX, USA) using the user-written runmlwin command (Leckie & Charlton, 2013), which calls the standalone multilevel modelling package MLwiN v3.01 (www.cmm.bristol.ac.uk/MLwiN/index.shtml). We note that MLwiN can also be called within R using the R2MLwiN package (Zhang, Parker, Charlton, Leckie, & Browne, 2016). For the analysis using the delta method, non-linear comparisons were estimated using the ‘nlcom’ command in Stata. Model equations and additional information are provided in the supplementary materials for further interpretation. To facilitate replication, annotated Stata code can be found here: https://github.com/kwongsiufung.

#### 2.3.1 Missing data

Missing data in the repeated measures data were handled using full information maximum likelihood (FIML), which assumes that data are missing at random (Little & Rubin, 2019). This means that once we account for the series of depressive symptoms scores and any covariate information that we do observe on a subject, the probability that they are missing in the data at any occasion does not depend on their unknown level of depressive symptoms at that occasion. Previous analysis has found that FIML is an appropriate method for handling missing data when examining trajectories of depressive symptoms (Kwong et al., 2019). Likewise, previous research has shown that there is no substantive change in the model estimates when removing individuals with more extreme missing data patterns (Kwong et al., 2019; Lopéz-Lopez et al., Forthcoming). We therefore include individuals in our analysis with at least one measurement of depressive symptoms in order to increase power to detect an association between the various trauma types and depressive symptoms.

## 3. Results

### 3.1. Sample characteristics

Participant demographics are described in Table 1. Additional descriptive statistics and reliability measures for the SMFQ are described in Supplementary Table 1. There were 6,711 individuals with data on the number of traumas, at least one measurement of depressive symptoms and all confounders. The interpretation of the results did not vary substantially with the inclusion of confounders, thus only the adjusted analysis are reported here. Unadjusted analyses are reported in Supplementary Tables 2 and 3.

**Table 1.** Participant demographics for the number of traumas

| | No Trauma | 1 Trauma | 2 Traumas | 3+ Traumas | $\chi^2, p$ |
| --- | --- | --- | --- | --- | --- |
| **Sex** |  |  |  |  |  |
| Males n (%) | 2,607 (45.70) | 1,166 (54.13) | 355 (49.44) | 141 (47.64) | $x^2 = 45.04, p < .001$ |
| Females n (%) | 3,907 (54.30) | 988 (45.87) | 363 (50.56) | 155 (52.36) |  |
| **Maternal Education** |  |  |  |  |  |
| A Level or Higher n (%) | 2,197 (40.60) | 838 (40.60) | 293 (42.80) | 125 (43.20) | $x^2 = 3.17, p = 0.79$ |
| O Level n (%) | 1,904 (35.20) | 737 (35.70) | 242 (35.40) | 97 (33.60) |  |
| < O Level n (%) | 1,311 (24.2) | 489 (23.70) | 149 (21.80) | 67 (23.20) |  |
| **Maternal Socioeconomic Status** |  |  |  |  |  |
| Professional/Managerial/Technical n (%) | 1,890 (40.80) | 704 (40.30) | 251 (42.80) | 95 (40.20) | $x^2 = 1.14, p = 0.77$ |
| Skilled non-manual or lower n (%) | 2,743 (59.20) | 1,043 (59.70) | 336 (57.20) | 141 (59.80) |  |
| **Parity** |  |  |  |  |  |
| First Born n (%) | 2,485 (45.20) | 996 (48.20) | 320 (46.90) | 117 (41.30) | $x^2 = 9.09, p = .169$ |
| Second Born n (%) | 1,998 (36.40) | 704 (34.10) | 233 (34.10) | 106 (37.50) |  |
| Third Born + n (%) | 1,012 (18.40) | 367 (17.70) | 130 (19.00) | 60 (21.20) |  |
| **Maternal Postnatal Depression** |  |  |  |  |  |
| No n (%) | 4,849 (92.60) | 1,790 (89.10) | 587 (87.30) | 288 (81.10) | $x^2 = 68.67, p < .001$ |
| Yes n (%) | 388 (7.40) | 219 (10.90) | 85 (12.70) | 53 (18.90) |  |

### 3.2. Direct model output vs alternative model parameterisation

Estimates from each trauma model can be parameterised in two ways. As the trauma variables were coded as dummy variables for both any trauma variable (coded as 0/1) and number of traumas (coded as 0/1/2/3+), the 0 coded dummy variable can be thought of as the baseline trajectory. Therefore, the effect of trauma (being coded as 1) or the number of traumas (coded as 1/2/3+) are ‘added’ onto the baseline trajectory to represent the effect of having any trauma/number of traumas. As shown in Tables 2 and 3, the direct model output shows the intercept and age term coefficients in the dummy variable regression format.

**Table 2.** Adjusted Associations Between Any Trauma (between 5-10 Years) and SMFQ Trajectories (n=6,664)

| Parameter | Direct Model Output |  |  | Alternative Parametrisation |  |  |
| --- | --- | --- | --- | --- | --- | --- |
|  | Estimate (95% CIs) | Std. Error | p-value | Estimate (95% CIs) | Std. Error | p-value |
| $\beta_0$ No Trauma Intercept | 5.17 [4.96, 5.38] | 0.11 | <.001 | 5.17 [4.96, 5.38] | 0.11 | <.001 |
| $\beta_1$ No Trauma x Age | 0.32 [0.28, 0.36] | 0.02 | <.001 | 0.32 [0.28, 0.36] | 0.02 | <.001 |
| $\beta_2$ No Trauma x Age<sup>2</sup> | -0.10 [-0.11, -0.08] | 0.01 | <.001 | -0.10 [-0.11, -0.08] | 0.01 | <.001 |
| $\beta_3$ No Trauma x Age<sup>3</sup> | -0.004 [0.01, -0.003] | 0.001 | <.001 | -0.004 [0.01, -0.003] | 0.001 | <.001 |
| $\beta_4$ No Trauma x Age<sup>4</sup> | 0.002 [0.001, 0.002] | 0.0001 | <.001 | 0.002 [0.001, 0.002] | 0.0001 | <.001 |
| $\beta_5$ Yes Trauma Intercept | 1.23 [0.97, 1.49] | 0.13 | <.001 | 6.40 [6.15, 6.65] | 0.13 | <.001 |
| $\beta_6$ Yes Trauma x Age | 0.06 [-0.01, 0.12] | 0.03 | 0.09 | 0.38 [0.33, 0.43] | 0.03 | 0.09 |
| $\beta_7$ Yes Trauma x Age<sup>2</sup> | -0.001 [-0.02, 0.02] | 0.01 | 0.95 | -0.10 [-0.11, 0.08] | 0.01 | 0.95 |
| $\beta_8$ Yes Trauma x Age<sup>3</sup> | -0.001 [-0.003, 0.003] | 0.001 | 0.11 | -0.006 [-0.007, 0.005] | 0.001 | 0.11 |
| $\beta_9$ Yes Trauma x Age<sup>4</sup> | 0.0001 [-0.0003, 0.0005] | 0.0002 | 0.69 | 0.002 [0.001, 0.002] | 0.0002 | 0.69 |
| Deviance | 189554.21 |  |  |  |  |  |

The no trauma variable should be viewed as the reference category as trauma was coded as a dummy variable (0/1). Thus, the intercept coefficient (and the coefficients for subsequent age terms) for an individual with trauma are the trauma intercept + yes trauma intercept (i.e., $\beta_0 + \beta_5$ ). The same applies for all age terms. The Alternative Parametrisation results can therefore be thought of as the calculated scores and therefore the $\beta_x$ refer to the direct model output only. The standard error and p-value correspond to the differences between the trauma estimates and the no trauma differences. The adjusted analysis included confounders (all entered as main effects) child sex, maternal postnatal depression, maternal education and social economic status at birth and parity. Intercepts are centred to age 16, the mean age of all assessments

**Table 3.**
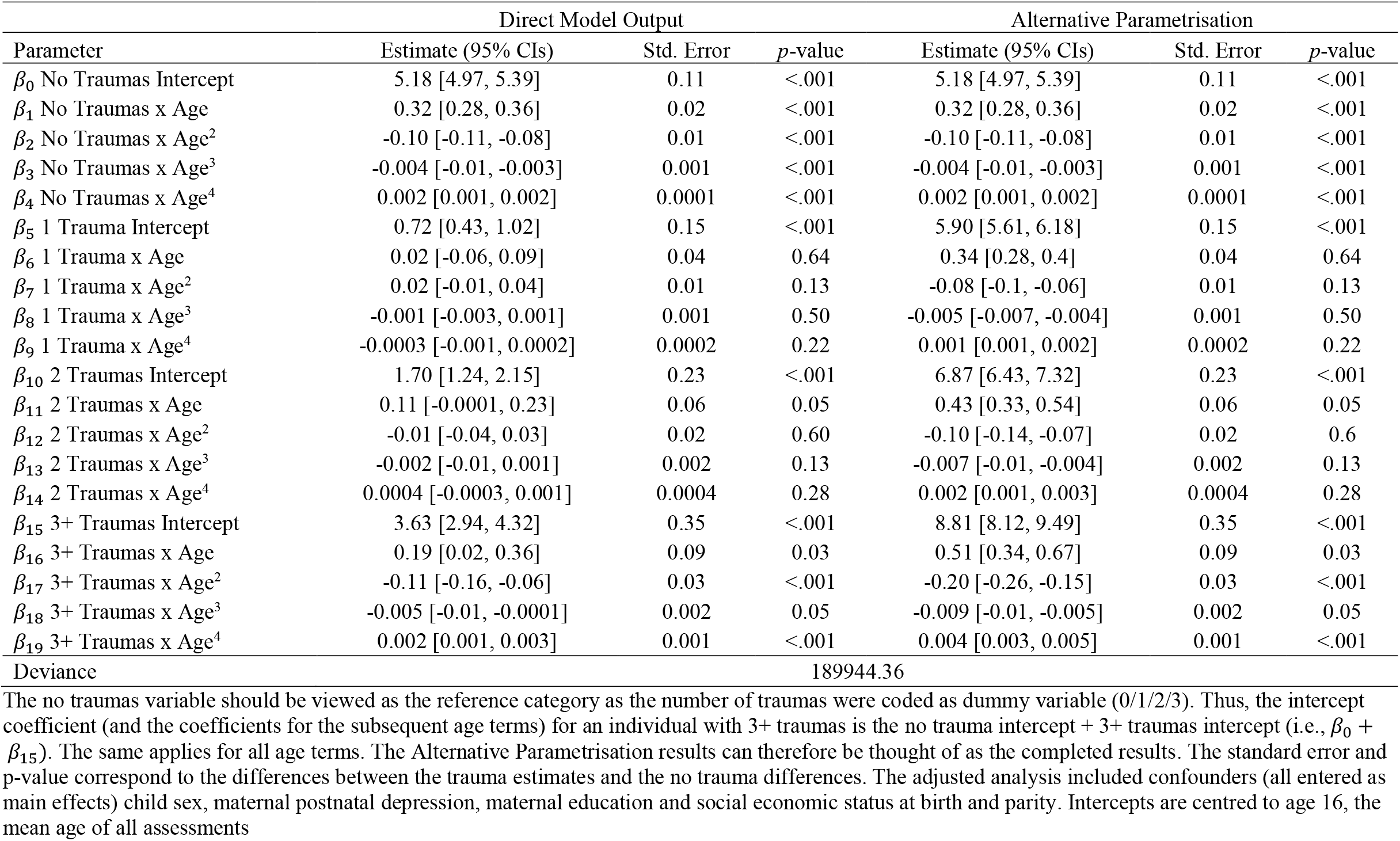
Adjusted Associations Between Number of Traumas (5-10 Years) and SMFQ Trajectories (n=6,711)

However, it is also possible to calculate an alternative parameterisation of the model in a more interpretable manner by showing the complete trajectory estimates for an individual with no trauma compared to an individual with trauma or the number of traumas. This is achieved by adding the coefficient of the trauma variable onto the coefficient of the reference variable which then calculates the intercept or age terms for each distinct population trajectory.

### 3.3. Association between any trauma and trajectories of depressive symptoms

As shown in Figure 1, individuals exposed to trauma (between ages 5-10) had worse trajectories of depressive symptoms compared to those who had not experienced trauma. Both trajectories increased throughout adolescence until the age of 18, where symptoms decreased. However, symptoms then began to rise again from the age of 22. The main differences between the two trajectories were characterised by individuals exposed to trauma having a higher intercept^1^ at age 16 (*β* = 6.4, SE= 0.13 [95% CI: 6.15, 6.65]) compared to non-trauma exposed individuals ((*β* = 5.17, SE= 0.11 [95% CI: 4.96, 5.38]; *p*^diff^ < .001). There was a weak association between individuals exposed to trauma and the linear age term (*β* = 0.38, SE= 0.03 [95% CI: 0.33, 0.43]), compared to non-trauma exposed individuals (*β* = 0.32, SE= 0.02 [95% CI: 0.28, 0.36]; *p*^diff^ = .09), which suggested that individuals with trauma may have higher linear growth. However, there was no association between the remaining age terms (e.g., quadratic, cubic and quartic age terms) and the non-trauma and trauma exposed individuals (*ps*^diff^ = .11), implying that overall growth was not substantially different between the two trajectories. Full estimates are shown in Table 2.

**Figure 1.**
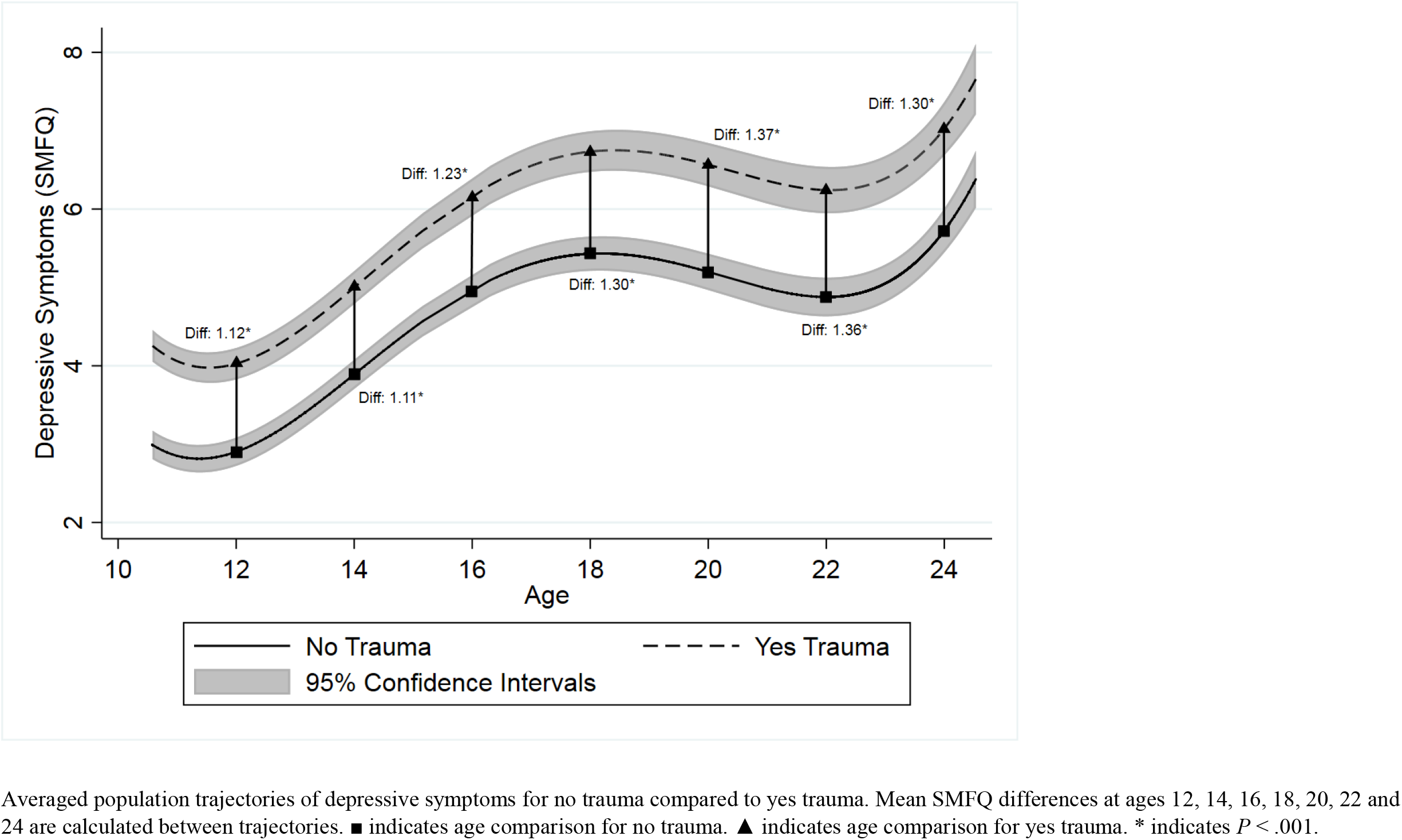
Adjusted Association Between Any Trauma (between 5-10 Years) and SMFQ Trajectories Averaged population trajectories of depressive symptoms for no trauma compared to yes trauma. Mean SMFQ differences at ages 12, 14, 16, 18, 20, 22 and 24 are calculated between trajectories. ■ indicates age comparison for no trauma. ▲ indicates age comparison for yes trauma. * indicates *P* < .001.

### 3.4. Association between the number of types of trauma and trajectories of depressive symptoms

As shown in Figure 2, there was an exposure-response relationship between the number of types of traumas (between ages 5-10) and later trajectories of depressive symptoms, with a tendency for more traumatic experiences in childhood to be associated with steeper trajectories of depressive symptoms. All four trajectories increased throughout adolescence and began to decrease from about the age of 18 and all trajectories again began to rise from the age of about 22. These differences between trajectories were mainly characterised by individuals who experienced no trauma having a lower intercept score at age 16 (*β* = 5.18, SE= 0.11 [95% CI: 4.97, 5.39]) compared to individuals with one trauma (*β* = 5.9, SE= 0.15 [95% CI: 5.61, 6.18]; *p*^diff^ < .001), two traumas (*β* = 6.87, SE= 0.23 [95% CI: 6.43, 7.32]; *p*^diff^ < .001) and three or more types of traumas (*β* = 8.81, SE= 0.35 [95% CI: 8.12, 9.49]; *p*^diff^ < .001). There was a weak association between individuals exposed to two traumas and the linear age term (*β* = 0.43, SE= 0.06 [95% CI: 0.33, 0.54]), compared to non-trauma exposed individuals (*β* = 0.32, SE= 0.02 [95% CI: 0.28, 0.36]; *p*^diff^ = .05), which suggested that individuals exposed to two traumatic experiences may have higher linear growth. There were no associations between the remaining age terms and the non-trauma, one trauma and two traumas exposed individuals (*ps*^diff^ = .13), suggesting that overall growth of the trajectories did not differ substantially between no trauma and the one/two traumas trajectories. There were also associations between individuals exposed to three or more types of traumatic experiences and the linear (*β* = 0.51, SE= 0.09 [95% CI: 0.34, 0.67]), quadratic (*β* = -0.2, SE= 0.03 [95% CI: -0.26, -0.15]), cubic (*β* = -0.009, SE= 0.002 [95% CI: -0.01, - 0.005]) and quartic age terms (*β* = 0.004, SE= 0.001 [95% CI: 0.003, 0.005]), compared to the linear (*β* = 0.32, SE= 0.02 [95% CI: 0.32, 0.36]; *p*^diff^ = .03), quadratic (*β* = -0.1, SE= 0.01 [95% CI: -0.11, -0.08]; *p*^diff^ = <.001), cubic (*β* = -0.004, SE= 0.001 [95% CI: -0.01, -0.003]; *p*^diff^ = .05) and quartic age terms (*β* = 0.002, SE= 0.0001 [95% CI: 0.001, 0.002]; *p*^diff^ = <.001) for those individuals who were not exposed to any trauma respectively. This suggested that individuals exposed to three or more types of traumas had less favourable trajectories that had a higher rate of change compared to the non-trauma exposed group. Full estimates are shown in Table 3.

**Figure 2.**
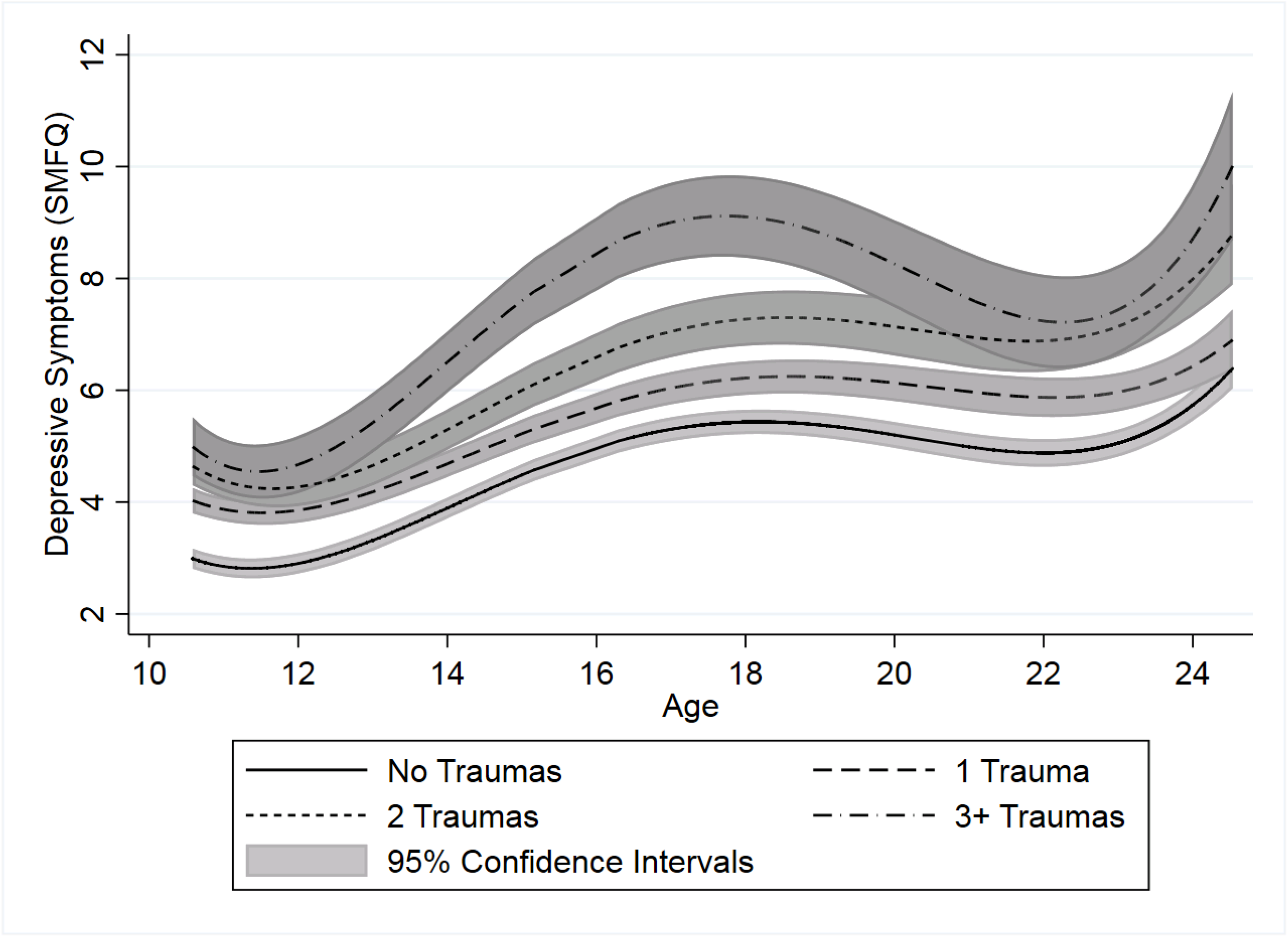
Adjusted Association Between Number of Trauma Types (between 5-10 Years) and SMFQ Trajectories

### 3.5. Comparing depressive symptom scores at different ages

To aid the interpretation of our analysis we compared predicted depressive symptoms scores at various ages throughout adolescence and young adulthood. We then examined whether these symptoms scores differed by the varying trajectories. To correct for multiple hypothesis testing, we used a Bonferroni adjustment of 0.001^2^ to estimate corrected main effects between these differences. As shown in Table 4, the predicted depressive symptoms scores at all ages were higher for individuals exposed to any trauma, compared to those who experienced no trauma (*ps*^diff^ = <.001). The largest difference between these two trajectories occurred at age 20 (*β* = 1.37, SE= 0.16, *p*^diff^ = <.001), which corresponded to a 5.27%^3^ difference in depressive symptoms.

**Table 4.**
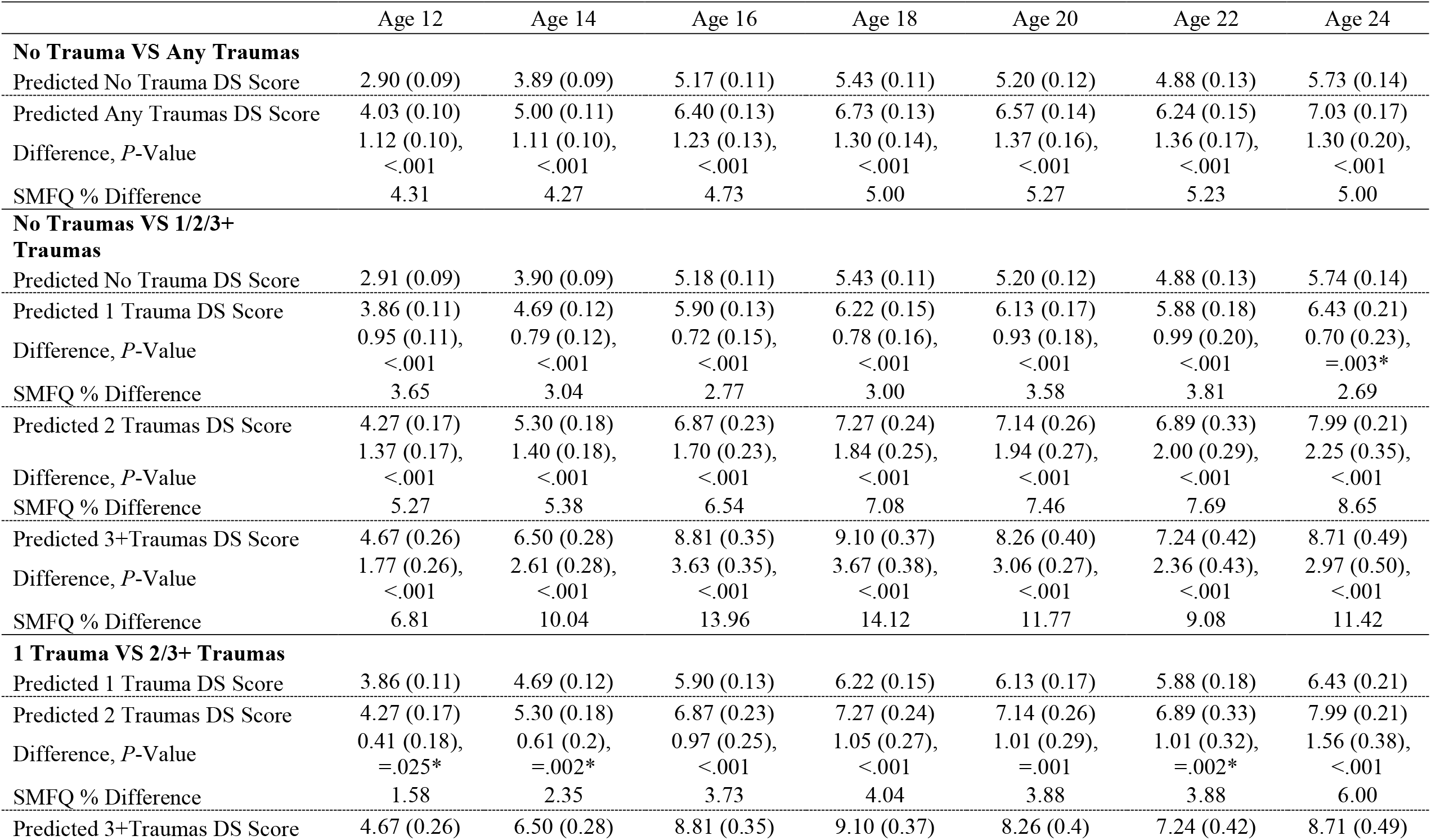
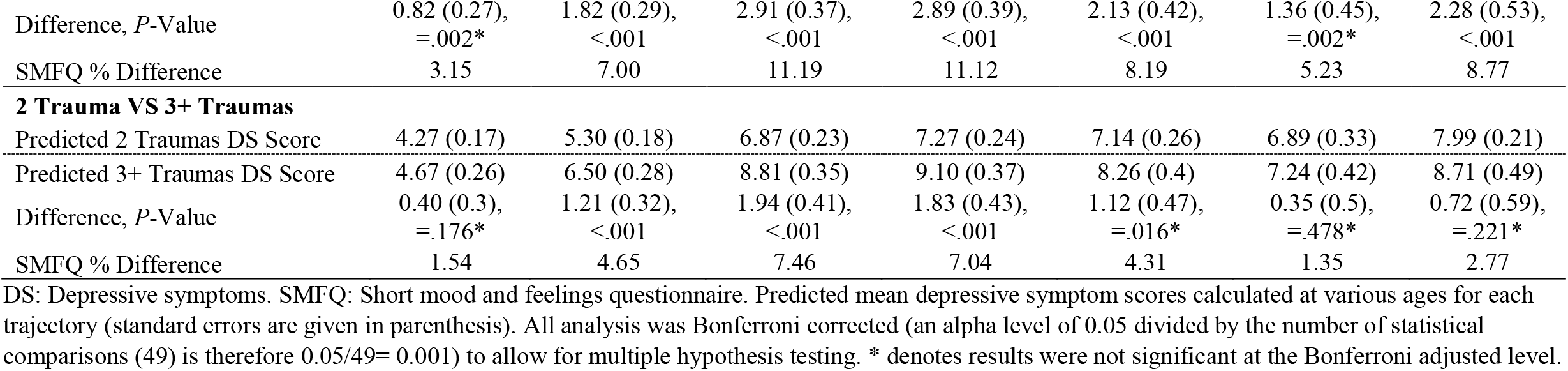
Comparisons Between Different Trauma Trajectories and Depressive Symptoms Scores at Various Ages

Results were comparable for the analysis on the number of traumas with individuals with more types of traumas having greater depressive symptoms score differences compared to those who had experienced no trauma. However, there was no difference in predicted depressive symptoms at age 24 between the no trauma trajectory and those with 1 trauma (*β* = 0.7, SE= 0.23, *p*^diff^ = .003 [Bonferroni adjusted]). Likewise, there were no differences between symptom scores for individuals in the one trauma trajectory, and those in the two traumas trajectory at ages 12, 14 and 22 (*ps*^diff^ > .0.002 [Bonferroni adjusted]), and the one trauma trajectory and the three or more trauma trajectory at ages 12 and 22 (*ps*^diff^ > .0.002 [Bonferroni adjusted]). There were no differences in symptoms scores between the two traumas trajectory and the three of more trauma trajectory at ages 12, 20, 22 and 24 (*ps*^diff^ > .0.016 [Bonferroni adjusted]). The largest difference between the trajectories occurred between the no trauma trajectory and three or more traumas trajectory at age 18 (*β* = 3.67, SE= 0.38, *p*^diff^ = <.001), which corresponded to a 14% difference in depressive symptoms.

## 4. Discussion

### 4.1. Main findings

We examined the association between childhood trauma and trajectories of depressive symptoms between the ages of 11 and 24 years. We also examined how the number of types of trauma exposure reported during mid-childhood may be associated with higher trajectories of depressive symptoms. One of our main goals was to convey these results in a more interpretable manner that might be beneficial for researchers, clinicians and policy makers. Previous research tends to just display the coefficients from a model, thus making it difficult to interpret results that are non-linear over time. We have provided a framework which easily transforms direct model output into more interpretable coefficients that can be compared across multiple stages of development. Figure 1 demonstrates the utility of this approach. The use of this framework quantifies how different population averaged trajectories vary between each other at varying times (i.e., how no trauma individuals differ from exposed to trauma individuals at age 12 or age 18 etc). We hope that future research will utilise this approach for other trajectory comparisons in the hope of developing and improving treatments and interventions.

Exposure to childhood trauma was strongly associated with a steeper trajectory of depressive symptoms. As well as having higher predicted depressive symptoms scores at age 16, predicted symptoms scores were also higher between the ages of 12 and 24. However, the growth or rate of change did not change substantively depending on exposure to trauma. Importantly, our analysis on the number of trajectories revealed an exposure-relationship response which demonstrated that a greater number of the types of traumatic experiences in childhood was associated with less favourable trajectories. Whilst the overall rate of change did not differ substantially between those not exposed to trauma and those who reported either one or two types of trauma exposure, individuals exposed to three or more trauma types (i.e., those who are polyvictimised) had a substantively higher rate of change compared to non-exposed individuals. Specifically, polyvictimised individuals had an increased rate of depressive symptoms throughout adolescence, which implies that depression was getting worse more rapidly compared to individuals who did not report exposure to trauma. Importantly, we also observed that depressive symptoms scores were higher at almost every two-year interval for the non-exposed individuals compared to those with exposure to one, two or three or more types of trauma. The greatest difference occurred between the no trauma trajectory and the three or more traumas trajectory at age 18 (corresponding to nearly a 4 point or 14% difference in depressive symptoms).

These findings support previous research examining the effects of trauma and negative life experiences on later trajectories of depressive symptoms (Barboza & Dominguez, 2017; Carlson & Oshri, 2018; Keane et al., 2018; Olino et al., 2010), and the number of stressful events on higher trajectories (Weeks et al., 2014). These findings are consistent with additional research that suggests that childhood trauma does impact on later depression (Copeland et al., 2018; Lewis et al., 2019) and efforts should be made to prevent traumatic experiences occurring in childhood and support those individuals who are exposed to trauma (Sara & Lappin, 2017). Our research highlights that those most at risk (i.e., polyvictimised in childhood) have substantially less favourable trajectories of depressive symptoms and efforts should be made to support this at-risk group, potentially through community and individual-level interventions to mitigate or prevent the risk of adverse mental health outcomes post-trauma and throughout adolescence.

There are several mechanisms, both psychological and biological, that have been considered to contribute to the pathway from exposure to traumatic experiences to the development of depression. Our findings of the association between exposure to trauma and the trajectory of depressive symptoms may inform theoretical frameworks of these mechanisms. Trauma in early life may have lasting effects on the risk of depression due to its impact during development, and as such reduces the ability to build resilience in adolescence which results in an increased risk of greater depressive symptoms later in life (Copeland et al., 2018; Dunn, Gilman, Willett, Slopen, & Molnar, 2012; Dunn, Nishimi, Gomez, Powers, & Bradley, 2018). Chronic exposure to childhood trauma is associated with repeated victimisation and psychological distress in adolescence (Dierkhising, Ford, Branson, Grasso, & Lee, 2019), and a range of negative functional outcomes in adulthood (Copeland et al., 2018). Previous research has also found that extreme stress during childhood and adolescence may have stronger or longer lasting effects on the HPA axis compared to trauma exposure in adulthood (Lupien, McEwen, Gunnar, & Heim, 2009), and is considered a key mediator in the relationship between trauma and negative mental health outcomes (Heim, Newport, Mletzko, Miller, & Nemeroff, 2008; Owens et al., 2018). Our findings for increased risk of depressive symptoms from exposure to trauma over the course of adolescence is consistent with evidence of sustained dopamine dysregulation from trauma exposure during early life.

### 4.2. Considerations

We have transformed the direct model output into two potentially more helpful frameworks which better highlight results and we consider this one of the key contributions of this work. The first is to transform the original model coefficients into a calculated set of alternative estimates: the “*Alternative Parametrisation*”. This simply adds the baseline intercept and age terms onto the additional groups giving a clearer idea of what depressive symptoms scores are at the intercept age and how they change over a one-year period. However, as the trajectories are non-linear and have multiple age terms, even this can be challenging to interpret, so the second approach is to create predicted depressive symptoms scores at various ages for different population averaged trajectories and to compare them with degrees of uncertainty. This takes into account the non-linearity of the age terms and represents an easy way to interpret the coefficients from the model. Both these methods improve interpretation from the direct model output and are important to highlight as it is important to know the coefficients for change over time, not just what the scores are at the intercept. Such analysis could be useful for individuals without statistical backgrounds, and it is much easier to interpret a predicted mean difference in symptoms at a particular age, rather than interpreting complex non-linear and contrasting coefficients (e.g., interpreting a positive linear age term but negative quadratic and positive cubic terms). Interpreting these coefficients in isolation is often not practical, but the framework presented in Table 4 and Figure 1 could aid in the interpretation of complex results into clinical practise and treatment.

However, there are certain considerations that must be made. Firstly, in this analysis, we chose to use multilevel growth-curve models. Alternative analysis could have used a growth mixture modelling or latent class approach to derive multiple sub-population trajectories and then compared depressive symptom scores at various ages between differing sub-population trajectories (i.e., differences in depression at age 12 between a stable low, increasing and decreasing trajectory). We chose to use a population-averaged approach with quartic polynomials in order to quantify the population averaged rate of change of depressive symptoms associated with exposure to varying degrees of trauma. However, higher order polynomials may constrict the data to take a quartic shape, thus giving estimates that are extrapolated beyond the data. This problem would also be present in the growth mixture and latent class approach. It could be possible to use other functions of time in place of higher order polynomials such as splines and fractional polynomials (Tilling, Macdonald-Wallis, Lawlor, Hughes, & Howe, 2014). However, extrapolation beyond the data is still as issue with these models. Thus, a safer option to account for extrapolation would be to refrain from computing estimates close to the tails of the trajectory. This would avoid calculating nonsensical estimates that are beyond the range of the date.

A second consideration regards the number of tests that could be run in the framework presented here. As shown in Table 4, we ran multiple comparisons between the no trauma and yes trauma trajectories, and with the no trauma and number of trauma trajectories, resulting in 49 statistical comparisons. Whilst it is important to compare these multiple ages, one concern is that by running multiple sets of analysis, the chances of obtaining a type one error increase (i.e., reporting a sample mean difference to be significant when there is not mean difference in the population: a chance result). Alternatively, such methods may also facilitate research that searches for “statistically significant” differences. This is not the point of such analysis and we advocate the use of Bonferroni adjusted comparisons, or false discovery rates (FDR) to minimise the impact of both scenarios.

### 4.3. Limitations and future research

Attrition plays a part in any longitudinal study. Missing data could be biasing our results. Previous analysis using similar methods has shown that when using multiple methods for dealing with missing data, the estimates do not change substantively (Lopez-Lopez et al., forthcoming; Kwong et al., 2019). To account for this missing data, we used full information maximum likelihood (FIML) which makes a missing at random (MAR) assumption.

However, even though we used this approach to account for missing data, we cannot fully rule out the effect of missing data on our results. One alternative approach would be to use multiple imputation where auxiliary variables can be introduced into the imputation model to make the MAR assumption more credible

Additionally, we only examined childhood trauma between the ages of 5 and 10 years old. There may be varying effects for traumas at earlier or later ages that could be investigated further. However, this was beyond the scope of this study and would likely require advanced modelling techniques, in order to approximately capture the time-varying element of childhood trauma. For instance, childhood trauma may cause depression, but there may also be reverse-causal effects from depressive symptoms on the risk of childhood trauma. Future models should take such modelling into account when examining the longitudinal relationship between childhood trauma and depression. Future research should also consider the timing and accumulation of exposures as these are important for later depression (Dunn, Soare, et al., 2018; Smith et al., 2016). For example, those who have exposure to multiple instances of trauma over consecutive periods of development may be at the greatest risk, but research has yet to explore this.

Also, many of the childhood trauma items were parent-reported, which may be subject to report bias. However, sensitivity analyses using this data has found little difference in parent-child reports for trauma (Croft et al., 2018). The measure of exposure frequency used in the study is based on the number of different types of trauma reported, which is only an approximate measure of how chronic trauma exposure is during the age period. There may be variation in the frequency of a single type of trauma exposure and its severity that this analysis is unable to account for. This study also assumes that there is an equal effect of each trauma type on the risk of depression, which may not be the case. These analyses were not the primary focus of this study, however, future research should look to explore the impact of different types of trauma and their associations with later depression as this may help identify at risk individuals, who could be targeted for interventions and treatment.

Previous research has also shown that sex differences in child trauma and depression may exist, and evidence suggests that stressful life events increase female trajectories of depressive symptoms, but not males (Adkins et al., 2009; Meadows, Brown, & Elder, 2006). However, the opposite effect has also been observed in another study (Fernandez Castelao & Kroner-Herwig, 2013). Sex differences were beyond the scope of this study as we were initially interested in presenting a model interpretation framework for trajectories analysis with a binary and then categorical variable. However, this methodology could be extended to run a three-way interaction between sex, exposure to trauma and varying trajectories of depressive symptoms.

In conclusion, exposure to childhood trauma was associated with less favourable trajectories of depressive symptoms. Further to this, there was an exposure-response relationship with the number of reported types of trauma, with individuals exposed to 3 or more childhood traumas having substantially higher trajectories of depressive symptoms. These results suggest that polyvictimised children should be considered at higher risk of depression during adolescence, which could inform intervention approaches. We have also provided an alternative model interpretation framework for examining and comparing trajectories of depressive symptoms, which shows depressive symptoms scores at different ages for different trajectories. This framework could be more interpretable for researchers, clinicians and policy workers, which could eventually aid in the translation of such findings into preventative interventions and treatments.

## Data Availability

Data access can be requested by submitting a proposal to the ALSPAC committee.

https://github.com/kwongsiufung

## Acknowledgements

We are extremely grateful to all the families who took part in this study, the midwives for their help in recruiting them, and the whole ALSPAC team, which includes interviewers, computer and laboratory technicians, clerical workers, research scientists, volunteers, managers, receptionists and nurses. The UK Medical Research Council and Wellcome (Grant Ref: 102215/2/13/2) and the University of Bristol provide core support for ALSPAC. A comprehensive list of grant funding is available on the ALSPAC website. This research was specifically funded by Wellcome (08426812/Z/07/Z), Wellcome and the MRC (076467/Z/05/Z; 092731; 092731; 092731), the MRC (MR/M006727/1), NIH (PD301198-SC101645). A.S.F.K is funded by an ESRC Advanced Quantitative Methods Studentship.

## Author Contributions

A.S.F.K had full access to all the data in the study and takes responsibility for the integrity of the data and the accuracy of the data analysis.

Study concept and design: A.S.F.K.

Acquisition, analysis, or interpretation of data: All authors.

Drafting of the manuscript: A.S.F.K.

Critical revision of the manuscript for important intellectual content: All authors.

Statistical analysis: A.S.F.K, J.M.M, G.L.

Administrative, technical, or material support: A.S.F.K.

Study supervision: G.L.

## Footnotes

1 Intercepts were centered to 16.53 years, the grand mean age. Centering the intercepts is common practice and allows for easier interpretation (Kwong et al., 2019).

2 The Bonferroni adjustment was calculated by assuming an alpha level of 0.05. We ran 49 statistical comparisons. Thus, the adjusted Bonferroni corrected level was calculated by 0.05/49 = 0.001. Thus we only declared comparisons with p-values less than 0.001 as significant at the 5% level.

3 The % difference in depressive symptoms was calculated by taking the predicted difference score (e.g., 1.37) and dividing it by the total maximum depressive symptoms score of 26 (1.37/26=5.27).

## References

Adkins, D. E., Wang, V., Dupre, M. E., van den Oord, E. J. C. J., & Elder, G. H. (2009). Structure and Stress: Trajectories of Depressive Symptoms across Adolescence and Young Adulthood. Social Forces, 88(1), 31–60.

Angold, A., Costello, E. J., Messer, S. C., & Pickles, A. (1995). Development of a short questionnaire for use in epidemiological studies of depression in children and adolescents. International Journal of Methods in Psychiatric Research, 5(4), 237–249.

Barboza, G. E., & Dominguez, S. (2017). Longitudinal growth of post-traumatic stress and depressive symptoms following a child maltreatment allegation: An examination of violence exposure, family risk and placement type. Children and Youth Services Review, 81, 368–378. doi:10.1016/j.childyouth.2017.08.029

Boyd, A., Golding, J., Macleod, J., Lawlor, D. A., Fraser, A., Henderson, J., … Davey Smith, G. (2013). Cohort Profile: the ‘children of the 90s’--the index offspring of the Avon Longitudinal Study of Parents and Children. Int J Epidemiol, 42(1), 111–127. doi:10.1093/ije/dys064

Carlson, M. W., & Oshri, A. (2018). Depressive Symptom Trajectories Among Sexually Abused Youth: Examining the Effects of Parental Perpetration and Age of Abuse Onset. Child Maltreat, 23(4), 387–398. doi:10.1177/1077559518779755

Copeland, W. E., Shanahan, L., Hinesley, J., Chan, R. F., Aberg, K. A., Fairbank, J. A., … Costello, E. J. (2018). Association of Childhood Trauma Exposure With Adult Psychiatric Disorders and Functional Outcomes. JAMA Netw Open, 1(7), e184493. doi:10.1001/jamanetworkopen.2018.4493

Croft, J., Heron, J., Teufel, C., Cannon, M., Wolke, D., Thompson, A., … Zammit, S. (2018). Association of Trauma Type, Age of Exposure, and Frequency in Childhood and Adolescence With Psychotic Experiences in Early Adulthood. JAMA Psychiatry. doi:10.1001/jamapsychiatry.2018.3155

Dierkhising, C. B., Ford, J. D., Branson, C., Grasso, D. J., & Lee, R. (2019). Developmental timing of polyvictimization: Continuity, change, and association with adverse outcomes in adolescence. Child Abuse Negl, 87, 40–50. doi:10.1016/j.chiabu.2018.07.022

Dunn, E. C., Gilman, S. E., Willett, J. B., Slopen, N. B., & Molnar, B. E. (2012). The impact of exposure to interpersonal violence on gender differences in adolescent-onset major depression: results from the National Comorbidity Survey Replication (NCS-R). Depress Anxiety, 29(5), 392–399. doi:10.1002/da.21916

Dunn, E. C., Nishimi, K., Gomez, S. H., Powers, A., & Bradley, B. (2018). Developmental timing of trauma exposure and emotion dysregulation in adulthood: Are there sensitive periods when trauma is most harmful? J Affect Disord, 227, 869–877. doi:10.1016/j.jad.2017.10.045

Dunn, E. C., Soare, T. W., Raffeld, M. R., Busso, D. S., Crawford, K. M., Davis, K. A., … Susser, E. S. (2018). What life course theoretical models best explain the relationship between exposure to childhood adversity and psychopathology symptoms: recency, accumulation, or sensitive periods? Psychol Med, 48(15), 2562–2572. doi:10.1017/S0033291718000181

Fernandez Castelao, C., & Kroner-Herwig, B. (2013). Different trajectories of depressive symptoms in children and adolescents: predictors and differences in girls and boys. J Youth Adolesc, 42(8), 1169–1182. doi:10.1007/s10964-012-9858-4

Ferro, M. A., Gorter, J. W., & Boyle, M. H. (2015a). Trajectories of depressive symptoms during the transition to young adulthood: the role of chronic illness. J Affect Disord, 174, 594–601. doi:10.1016/j.jad.2014.12.014

Ferro, M. A., Gorter, J. W., & Boyle, M. H. (2015b). Trajectories of Depressive Symptoms in Canadian Emerging Adults. American Journal of Public Health, 105(11).

Fraser, A., Macdonald-Wallis, C., Tilling, K., Boyd, A., Golding, J., Davey Smith, G., … Lawlor, D. A. (2013). Cohort Profile: the Avon Longitudinal Study of Parents and Children: ALSPAC mothers cohort. Int J Epidemiol, 42(1), 97–110. doi:10.1093/ije/dys066

Ge, X., Lorenz, F. O., Conger, R. D., Elder, G. H., & Simons, R. L. (1994). Trajectories of Stressful Life Events and Depressive Symptoms During Adolescence. Developmental Psychology, 30(4), 467–483.

Ge, X., Natsuaki, M. N., & Conger, R. D. (2006). Trajectories of depressive symptoms and stressful life events among male and female adolescents in divorced and nondivorced families. Dev Psychopathol, 18(1), 253–273. doi:10.1017/S0954579406060147

Hedeker, D., & Gibbons, R. D. (2006). Longitudinal Data Analysis. Hoboken, New Jersey: John Wiley & Sons, Inc.

Heim, C., Newport, D. J., Mletzko, T., Miller, A. H., & Nemeroff, C. B. (2008). The link between childhood trauma and depression: insights from HPA axis studies in humans. Psychoneuroendocrinology, 33(6), 693–710. doi:10.1016/j.psyneuen.2008.03.008

Houtepen, L. C., Heron, J., Suderman, M. J., Tilling, K., & Howe, L. D. (2018). Adverse childhood experiences in the children of the Avon Longitudinal Study of Parents and Children (ALSPAC). Wellcome Open Res, 3, 106. doi:10.12688/wellcomeopenres.14716.1

Keane, C. A., Magee, C. A., & Kelly, P. J. (2018). Trajectories of Psychological Distress in Australians Living in Urban Poverty: The Impact of Interpersonal Trauma. J Trauma Stress, 31(3), 362–372. doi:10.1002/jts.22297

Kingsbury, M., Weeks, M., MacKinnon, N., Evans, J., Mahedy, L., Dykxhoorn, J., & Colman, I. (2016). Stressful Life Events During Pregnancy and Offspring Depression: Evidence From a Prospective Cohort Study. J Am Acad Child Adolesc Psychiatry, 55(8), 709–716 e702. doi:10.1016/j.jaac.2016.05.014

Kwong, A. S. F., Manley, D., Timpson, N. J., Pearson, R. M., Heron, J., Sallis, H., … Leckie, G. (2019). Identifying Critical Points of Trajectories of Depressive Symptoms from Childhood to Young Adulthood. J Youth Adolesc, 48, 815–827. doi:10.1007/s10964-018-0976-5

Leckie, G., & Charlton, C. (2013). runmlwin: A Program to Run the MLwiN Multilvel Modeling Software from within Stata. Journal of Statistical Software, 52(11).

Lee, T. K., Wickrama, K. A. S., Kwon, J. A., Lorenz, F. O., & Oshri, A. (2017). Antecedents of transition patterns of depressive symptom trajectories from adolescence to young adulthood. Br J Dev Psychol, 35(4), 498–515. doi:10.1111/bjdp.12189

Lewis, S. J., Arseneault, L., Caspi, A., Fisher, H. L., Matthews, T., Moffitt, T. E., … Danese, A. (2019). The epidemiology of trauma and post-traumatic stress disorder in a representative cohort of young people in England and Wales. The Lancet Psychiatry, 6(3), 247–256. doi:10.1016/s2215-0366(19)30031-8

Little, R. J. A., & Rubin, D. B. (2019). Statistical analysis with missing data (3rd Edition ed.). Hoboken, NJ: John Wiley & Sons.

Lopéz-Lopez, J. A., Kwong, A. S. F., Pearson, R. M., Tilling, K. M., Fazel, M. S., Washbrook, L., … Hammerton, G. (Forthcoming). Trajectories of depressive symptoms through adolescence and associations with education and employment: a growth mixture modelling approach.

Lupien, S. J., McEwen, B. S., Gunnar, M. R., & Heim, C. (2009). Effects of stress throughout the lifespan on the brain, behaviour and cognition. Nat Rev Neurosci, 10(6), 434–445. doi:10.1038/nrn2639

Mahedy, L., Hammerton, G., Teyhan, A., Edwards, A. C., Kendler, K. S., Moore, S. C., … Heron, J. (2017). Parental alcohol use and risk of behavioral and emotional problems in offspring. PLoS One, 12(6), e0178862. doi:10.1371/journal.pone.0178862

Mandy, W., Pellicano, L., St Pourcain, B., Skuse, D., & Heron, J. (2018). The development of autistic social traits across childhood and adolescence in males and females. J Child Psychol Psychiatry. doi:10.1111/jcpp.12913

Marmorstein, N. R. (2009). Longitudinal associations between alcohol problems and depressive symptoms: early adolescence through early adulthood. Alcohol Clin Exp Res, 33(1), 49–59. doi:10.1111/j.1530-0277.2008.00810.x

Mazza, J. J., Fleming, C. B., Abbott, R. D., Haggerty, K. P., & Catalano, R. F. (2010). Identifying trajectories of adolescents’ depressive phenomena: an examination of early risk factors. J Youth Adolesc, 39(6), 579–593. doi:10.1007/s10964-009-9406-z

Meadows, S. O., Brown, J. S., & Elder, G. H. (2006). Depressive Symptoms, Stress, and Support: Gendered Trajectories From Adolescence to Young Adulthood. Journal of Youth and Adolescence, 35(1), 89–99. doi:10.1007/s10964-005-9021-6

Mumford, E. A., Liu, W., Hair, E. C., & Yu, T. C. (2013). Concurrent trajectories of BMI and mental health patterns in emerging adulthood. Soc Sci Med, 98, 1–7. doi:10.1016/j.socscimed.2013.08.036

Musliner, K. L., Munk-Olsen, T., Eaton, W. W., & Zandi, P. P. (2016). Heterogeneity in long-term trajectories of depressive symptoms: Patterns, predictors and outcomes. J Affect Disord, 192, 199–211. doi:10.1016/j.jad.2015.12.030

Muthen, B., & Muthen, L. K. (2000). Integrating Person-Centered and Variable-Centered Analyses: Growth Mixture Modeling With Latent Trajectory Classes. ALCOHOLISM: CLINICAL AND EXPERIMENTAL RESEARCH, 24(6).

Olino, T. M., Klein, D. N., Lewinsohn, P. M., Rohde, P., & Seeley, J. R. (2010). Latent trajectory classes of depressive and anxiety disorders from adolescence to adulthood: descriptions of classes and associations with risk factors. Compr Psychiatry, 51(3), 224–235. doi:10.1016/j.comppsych.2009.07.002

Owens, S. A., Helms, S. W., Rudolph, K. D., Hastings, P. D., Nock, M. K., & Prinstein, M. J. (2018). Interpersonal Stress Severity Longitudinally Predicts Adolescent Girls’ Depressive Symptoms: the Moderating Role of Subjective and HPA Axis Stress Responses. J Abnorm Child Psychol. doi:10.1007/s10802-018-0483-x

Raudenbush, S. W., & Bryk, A. S. (2002). Hierarchical Linear Models: Applications and Data Analysis Methods (Second Edition ed.). Thousand Oaks, California: Sage Publications, Inc.

Rawana, J. S., & Morgan, A. S. (2014). Trajectories of depressive symptoms from adolescence to young adulthood: the role of self-esteem and body-related predictors. J Youth Adolesc, 43(4), 597–611. doi:10.1007/s10964-013-9995-4

Rice, F., Riglin, L., Thapar, A. K., Heron, J., Anney, R., O’Donovan, M. C., & Thapar, A. (2018). Characterizing Developmental Trajectories and the Role of Neuropsychiatric Genetic Risk Variants in Early-Onset Depression. JAMA Psychiatry. doi:10.1001/jamapsychiatry.2018.3338

Sallinen, M., Rönkä, A., Kinnunen, U., & Kokko, K. (2016). Trajectories of depressive mood in adolescents: Does parental work or parent-adolescent relationship matter? A follow-up study through junior high school in Finland. International Journal of Behavioral Development, 31(2), 181–190. doi:10.1177/0165025407074631

Sara, G., & Lappin, J. (2017). Childhood trauma: psychiatry’s greatest public health challenge? The Lancet Public Health, 2(7), e300–e301. doi:10.1016/s2468-2667(17)30104-4

Schubert, K. O., Clark, S. R., Van, L. K., Collinson, J. L., & Baune, B. T. (2017). Depressive symptom trajectories in late adolescence and early adulthood: A systematic review. Aust N Z J Psychiatry, 51(5), 477–499. doi:10.1177/0004867417700274

Shore, L., Toumbourou, J. W., Lewis, A. J., & Kremer, P. (2018). Review: Longitudinal trajectories of child and adolescent depressive symptoms and their predictors - a systematic review and meta-analysis. Child and Adolescent Mental Health, 23(2), 107–120. doi:10.1111/camh.12220

Singer, J. D., & Willett, J. B. (2003). Applied Longitudinal Data Analysis: Modelling Change and Event Occurence. New York: Oxford University Press.

Smith, A. D., Hardy, R., Heron, J., Joinson, C. J., Lawlor, D. A., Macdonald-Wallis, C., & Tilling, K. (2016). A structured approach to hypotheses involving continuous exposures over the life course. Int J Epidemiol, 45(4), 1271–1279. doi:10.1093/ije/dyw164

Stoolmiller, M., Kim, H. K., & Capaldi, D. M. (2005). The course of depressive symptoms in men from early adolescence to young adulthood: identifying latent trajectories and early predictors. J Abnorm Psychol, 114(3), 331–345. doi:10.1037/0021-843X.114.3.331

Sutin, A. R., Terracciano, A., Milaneschi, Y., An, Y., Ferrucci, L., & Zonderman, A. B. (2013). The trajectory of depressive symptoms across the adult life span. JAMA Psychiatry, 70(8), 803–811. doi:10.1001/jamapsychiatry.2013.193

Thapar, A., Collinshaw, S., Pine, D. S., & Thapar, A. J. (2012). Depression in adolescence. Lancet, 379, 1056–1067.

Thapar, A., & McGuffin, P. (1998). Validity of the shortened Mood and Feelings Questionnaire in a community sample of children and adolescents: a preliminary research note. Psychiatry Research, 81, 259–268.

Tilling, K., Macdonald-Wallis, C., Lawlor, D. A., Hughes, R. A., & Howe, L. D. (2014). Modelling childhood growth using fractional polynomials and linear splines. Ann Nutr Metab, 65(2-3), 129–138. doi:10.1159/000362695

Turner, N., Joinson, C., Peters, T.J., Wiles, N., & Lewis, G. (2014). Validity of the Short Mood and Feelings Questionnaire in Late Adolescence. Psychological Assessment. doi:10.1037/a0036572.supp

Weeks, M., Cairney, J., Wild, T. C., Ploubidis, G. B., Naicker, K., & Colman, I. (2014). Early-life predictors of internalizing symptom trajectories in Canadian children. Depress Anxiety, 31(7), 608–616. doi:10.1002/da.22235

Whalen, D. J., Luby, J. L., Tilman, R., Mike, A., Barch, D., & Belden, A. C. (2016). Latent class profiles of depressive symptoms from early to middle childhood: predictors, outcomes, and gender effects. J Child Psychol Psychiatry, 57(7), 794–804. doi:10.1111/jcpp.12518

Wiesner, M., & Kim, H. K. (2006). Co-occurring delinquency and depressive symptoms of adolescent boys and girls: a dual trajectory modeling approach. Dev Psychol, 42(6), 1220–1235. doi:10.1037/0012-1649.42.6.1220

Yaroslavsky, I., Pettit, J. W., Lewinsohn, P. M., Seeley, J. R., & Roberts, R. E. (2013). Heterogeneous trajectories of depressive symptoms: adolescent predictors and adult outcomes. J Affect Disord, 148(2-3), 391–399. doi:10.1016/j.jad.2012.06.028

Zhang, Z., Parker, R., Charlton, C., Leckie, G., & Browne, W. J. (2016). R2MLwiN - A program to run the MLwiN multilevel modelling software from within R. Journal of Statistical Software, 72(10), 1–43. doi:DOI: 10.18637/jss.v072.i10

